# Assessing the Mediterranean diet adherence during pregnancy: practical considerations based on the associations with cardiometabolic risk

**DOI:** 10.1101/2022.09.20.22280165

**Authors:** Marta Flor-Alemany, Jairo H. Migueles, Pedro Acosta-Manzano, Nuria Marín-Jiménez, Laura Baena-García, Virginia A Aparicio

## Abstract

**Objective:** The aim of the present study was to provide practical considerations for assessing MD adherence during pregnancy based on the association with cardiometabolic risk.

**Study design:** Longitudinal study

**Main outcome measures:** A food frequency questionnaire was fulfilled by 152 pregnant women at the 16^th^ gestational week (g.w.). We calculated the Mediterranean Food Pattern (MFP), the MD Scale (MDScale), the Short MD questionnaire (SMDQ), the MD Score (MedDietScore), and the MD scale for pregnant women (MDS-P). The cardiometabolic risk score consisted of pre-pregnancy body mass index, blood pressure, glucose, triglycerides, and high-density lipoprotein-cholesterol (at 16^th^ and 34^th^ g.w.).

**Results:** Multiple linear regression models showed that the MFP, the MedDietScore, and the SMDQ were associated with lower cardiometabolic risk at the 16^th^ and 34^th^ g.w. (β’s: - 0.193 to -0.415, all *p*<0.05); and the MDS-P at the 34^th^ g.w. (β=-0.349, *p*<0.01). A comparison of these models with the *J* test showed that the MFP and the MedDietScore outperformed the SMDQ at the 16^th^ g.w. (*p’s*<0.05); while the MedDietScore outperformed the SMDQ, MFP, and MDS-P (*p’s*<0.05) at the 34^th^ g.w. Receiver-Operating-Characteristic-derived thresholds for the MFP, MedDietScore and MDS-P indices were 21, 30, and 6 points, respectively, to identify women with high cardiometabolic risk.

**Conclusion:** The MFP and MedDietScore are recommended to assess MD adherence during pregnancy, as these showed the strongest associations with cardiometabolic risk. Our validated thresholds might assist in the detection of poor dietary patterns during pregnancy.

## INTRODUCTION

The gestational period is a crucial time of growth, development, and physiological changes for the mother and child[1]. Diet during pregnancy is key for the health status of both the mother and the new-born[2,3]. Although previous evidence is based on a single or a few food items or nutrients[3], the diet should be considered as the combination of the nutritive and non-nutritive components[4]. Dietary patterns can comprehensively and meaningfully assess the relation between the diet quality and pregnancy outcomes[2]. The Mediterranean diet (MD) is one of the healthiest dietary patterns recognized in Europe[5].

MD in pregnancy lowers the maternal cardiometabolic risk[6], the risk for excessive gestational weight gain, gestational diabetes[7], and preterm delivery[8], and the offspring insulin resistance[9] and adiposity[10]. However, most of the MD indices used in pregnancy are not specifically validated in pregnant women[11]. The dietary habits are affected during pregnancy (e.g., avoid alcohol) and require specific assessment tools. Thus far, there is only a MD index that has been adapted for pregnant women[12], yet the cut-off points to interpret its score were not adapted. As a result, it is challenging to assess the MD adherence during pregnancy[3,10].

Assessing MD adherence in pregnancy is especially relevant since recent studies suggest that pregnant women are drifting away from the MD-like pattern[13,14]. This study aimed to provide practical considerations on what Mediterranean diet (MD) indices are useful to assess MD adherence during gestation based on their association with a clustered cardiometabolic risk at the 16^th^ and 34^th^ gestational weeks (g.w.).

## METHODS

### Study design and participants

This longitudinal study forms part of the GEStation and FITness (GESTAFIT) project[15]. The GESTAFIT delivered an exercise intervention to pregnant women (more details elsewhere[15]). From the 384 pregnant women assessed for eligibility, 159 met the eligibility criteria (**Supplementary Table S1)** and signed a written informed consent. Among them, 152 had valid data in sociodemographic and clinical characteristics and MD adherence (**Supplementary Figure S1**). All procedures were approved by the Ethics Committee on Clinical Research of Granada, Regional Government of Andalusia, Spain (code: GESFIT-0448-N-15).

### Maternal anthropometry and body composition

Pre-pregnancy body weight was self-reported as the women were enrolled in this study in the 12^th^ g.w. Although measured weight is preferable, self-report is a cost-effective and practical measurement approach that shows very good concordance with measured body weight[16]. Height was measured using a stadiometer (Seca 22, Hamburg, Germany) at the 16^th^ g.w. Pre-pregnancy body mass index (BMI) was calculated as pre-pregnancy weight (kg) divided by squared height (m^2^).

### Cardiometabolic health markers

Blood pressure was assessed twice (with 2 minutes between trials) with an upper-arm blood pressure monitor (Omron M6, Omron Health Care Europe B.V. Hoolddorp, The Netherlands) with the person seated. The lowest value was selected for the analysis.Blood samples were collected after all-night fasting. Venous blood samples (5mL) were collected in serum tubes. After 30 minutes at room temperature (to allow the sample to clot), samples were centrifuged at 1750 rpm for 10 min at 4ºC in a refrigerated centrifuge (GS-6R Beckman Coulter, Brea, CA, USA) to obtain serum. Glucose, triglycerides, and high-density lipoprotein-cholesterol (HDL-C) were assessed with standard procedures using an auto-analyzer (AU5822 Clinical Chemistry Analyzer, Beckman Coulter, Brea, CA, USA).

### Cardiometabolic risk

A clustered cardiometabolic risk score was created as previously described by Lei et al. [17] from the Z-scores for pre-pregnancy BMI, mean blood pressure [defined as (SBP+DBP)/2], serum fasting glucose, triglycerides and HDL-C at the 16^th^ and 34^th^ g.w. The HDL-C z-score was multiplied by -1 prior to be averaged with the rest of factors. Higher clustered cardiometabolic status indicates greater cardiometabolic risk.

### Dietary assessment

A food frequency questionnaire (FFQ) validated in Spanish non-pregnant adults was employed[18]. The FFQ[18] consisted of a list of foods, portion sizes, and food groups adapted to the Spanish population[19]. Participants were asked about the frequency of consumption of the different foods, and additionally about meal and snack patterning with two extra questions. Energy (Kcal), macro, and micronutrient intake were calculated using the Evalfinut software[19].

We calculated five MD adherence indices from the FFQ data[18]: The Mediterranean Food Pattern (MFP)[20], the Mediterranean Diet Scale (MDScale)[21], the Short Mediterranean Diet questionnaire (SMDQ)[22], the Mediterranean Diet Score (MedDietScore)[23], and the Mediterranean Diet scale for pregnant women (MDS-P)[24]. The indices were calculated following the recommendations in the validation studies (further detailed in **Table 1**). Higher scores indicate higher diet quality. A moderate alcohol intake, also typical of the MD, was not considered for calculating these indices in this group of women These dietary indices have been previously associated with better cardiometabolic health in pregnant[3,24–28] or non-pregnant populations[22,23,29–32]. The components included in each index are detailed in **Supplementary Table S2**.

**Table 1.** Definition and references for the MD indices calculated in this study.

| Index | Definition | Original Range | Range adapted to pregnant women | Reference |
| --- | --- | --- | --- | --- |
| MFP | Quintile-based sum score in g/day of olive oil, fiber, fruits, vegetables, fish, cereals, meat and subproducts. Intake of each of the indicated groups in grams per day was calculated. The distribution was calculated within the study sample and each participant was assigned a score from 1 to 5 depending on the quintile of intake. The elements were combined by adding the quintile values. | 8-40 | 7-35 | [20] |
| MDScale | It consists of eight components in g/day (cereals, legumes, fruits and nuts, fish, high-fat dairy, meat, alcohol, and ratio of monounsaturated lipids to saturated lipids). Participants whose consumption of meat and dairy was the median were assigned a value of 1, and participants whose consumption was at or above the median were assigned a value of 0. The other components were assumed to be beneficial and were scored on a reverse scale. | 0-9 | 0-8 | [21] |
| SMDQ | A specific frequency score was assigned to each food component (1 attributable point) when food consumption met the established criteria for nine elements including olive oil, fruit, vegetables or salad, fruits and vegetables, legumes, fish, wine, meat, and cereals (wholegrain bread, white bread, and white rice). | 0-9 | 0-8 | [22] |
| MedDietScore | It consists of eleven variables (wholegrain cereals, potatoes, fruits, vegetables, pulses, fish, olive oil, red wine, red meat and subproducts, poultry and whole dairy products) ranging from 0 to 5 according to their position in the Mediterranean Diet pyramid. | 0-55 | 0-50 | [23] |
| MDS-P | It consists of eleven components including vegetables, fruits and nuts, legumes, cereals, fish, dairy, meat, the ratio of monosaturated to saturated fatty acids, folic acid intake, calcium intake, and iron intake. A high intake (above participants' median intake) of vegetables, fruits and nuts, pulses, cereals, fish, high monounsaturated to saturated fatty acids, and low intake (below the participants' median intake) of meat and dairy products scoring 1 point per component. The intake of folic acid, iron, and calcium was scored in relation to the Spanish recommended daily intake[19], scoring 1 point if the intake of the micronutrients was greater or equal to two-thirds of the recommended daily intake or if the women were taking nutritional supplements. | 0-11 | 0-11 | [24] |

MDScale, Mediterranean Diet Scale; MedDietScore, Mediterranean Diet Score; MDS-P, Mediterranean diet scale for pregnant women; MFP, Mediterranean Food Pattern; SMDQ, Short Mediterranean Diet questionnaire.

### Statistical analysis

Descriptive data were summarized as mean (standard deviation, SD) or frequency (%) as appropriate. Linear regression models adjusted for maternal age, smoking habit, and number of children, were used to explore the cross-sectional associations between MD adherence and clustered cardiometabolic risk at the 16^th^ g.w. (n=119). The longitudinal associations including the clustered cardiometabolic risk at the 34^th^ g.w. (n=107) were additionally adjusted for the exercise intervention. We used the *J* test[33] to compare the regression models including different MD indices. Likewise, multiple linear regression analysis was performed for the association of MD indices with the individual cardiometabolic markers (i.e., pre-pregnancy BMI, SBP, DBP, glucose, triglycerides, and HDL-C) adjusting for the aforementioned confounders. We explored the interaction between age (0= below 33 years old and 1= above 33 years old) and the MD adherence (with all MD indices) on clustered cardiometabolic risk during pregnancy. Since the clustered cardiometabolic risk*MD adherence interaction term was not significant (all *p*’s>0.2) we decided not to conduct separate models for women according to age categories.

We investigated the classification capacity to detect high cardiometabolic risk with receiver Operating Characteristic (ROC) curves at the 16^th^ and 34^th^ g.w. As such, women in the tertile 3 of the clustered cardiometabolic risk score were considered to be at high risk. The area under the curve (AUC) was obtained from the sensitivity versus specificity curves as a measure of diagnostic accuracy for each index, and 95% confidence intervals (95% CI) for the AUCs were derived[34]. AUC values of 0.90 were considered excellent; 0.80–0.89, good; 0.70–0.79, fair; and <0.70, poor[35]. Then, thresholds were developed, seeking to maximize both sensitivity (i.e., true positives) and specificity (i.e., true negatives). As such, the closest threshold to the perfect sensitivity and specificity was identified, i.e., the minimum value in equation[36]: (1-sensitivities)^2^ + (1 - specificities)^2^. The percentage of agreement and the relationship in the classification across indices was assessed with Kappa coefficients over the ROC-derived thresholds.[37]

Linear regression models were conducted using the Statistical Package for Social Sciences (IBM SPSS Statistics for Windows, version 22.0, Armonk, NY), the *J* test and the ROC analyses were conducted with the lmtest and the pROC R packages (v.4.1.1)[38]. The level of significance was set at *p*≤0.05.

## RESULTS

Of the 152 pregnant women (33.9±4.6 years) included, 119 had valid cardiometabolic data at the 16^th^ g.w., and 107 at the 34^th^ g.w. **(Supplementary Figure S1)**. The sociodemographic and clinical characteristics are shown in **Table 2**.

**Table 2.**
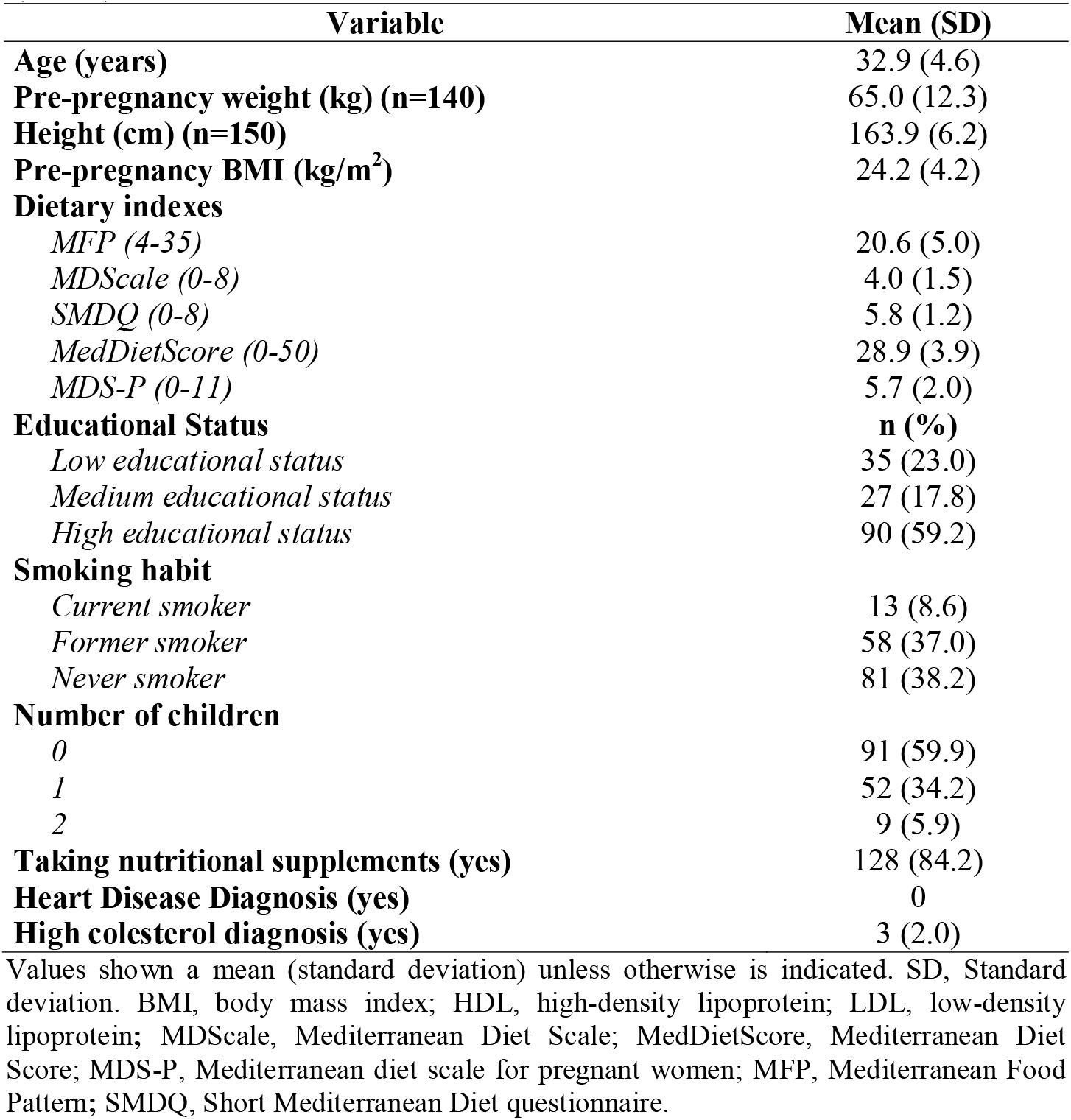
Sociodemographic and clinical characteristics of the study participants (n=152).

The cross-sectional and longitudinal associations of the MD indices with the clustered cardiometabolic risk (Z-score) are shown in **Figure 1**. Greater adherence to the MFP, SMDQ, and MedDietScore were significantly associated with lower clustered cardiometabolic risk at the 16^th^ g.w. and 34^th^ g.w. (β’s ranging from -0.193 to -0.415, *p’s*<0.05). In addition, a greater MDS-P score was associated with lower clustered cardiometabolic risk at the 34^th^ g.w. (β=-0.349, *p*<0.01). The *J* test analyses showed no significant differences in the models including the MFP and the MedDietScore at the 16^th^ g.w. (*p*=0.256), while the SMDQ showed a lower performance than the MFP (*p*=0.05) and the MedDietScore (*p*=0.027). At the 34^th^ g.w., the models including the MFP and the MDS-P were not statistically different (*p*=0.138), while the MedDietScore outperformed the MFP (*p*=0.003), the SMDQ (*p*<0.001) and the MDS-P (*p*<0.001).

**Figure 1.**
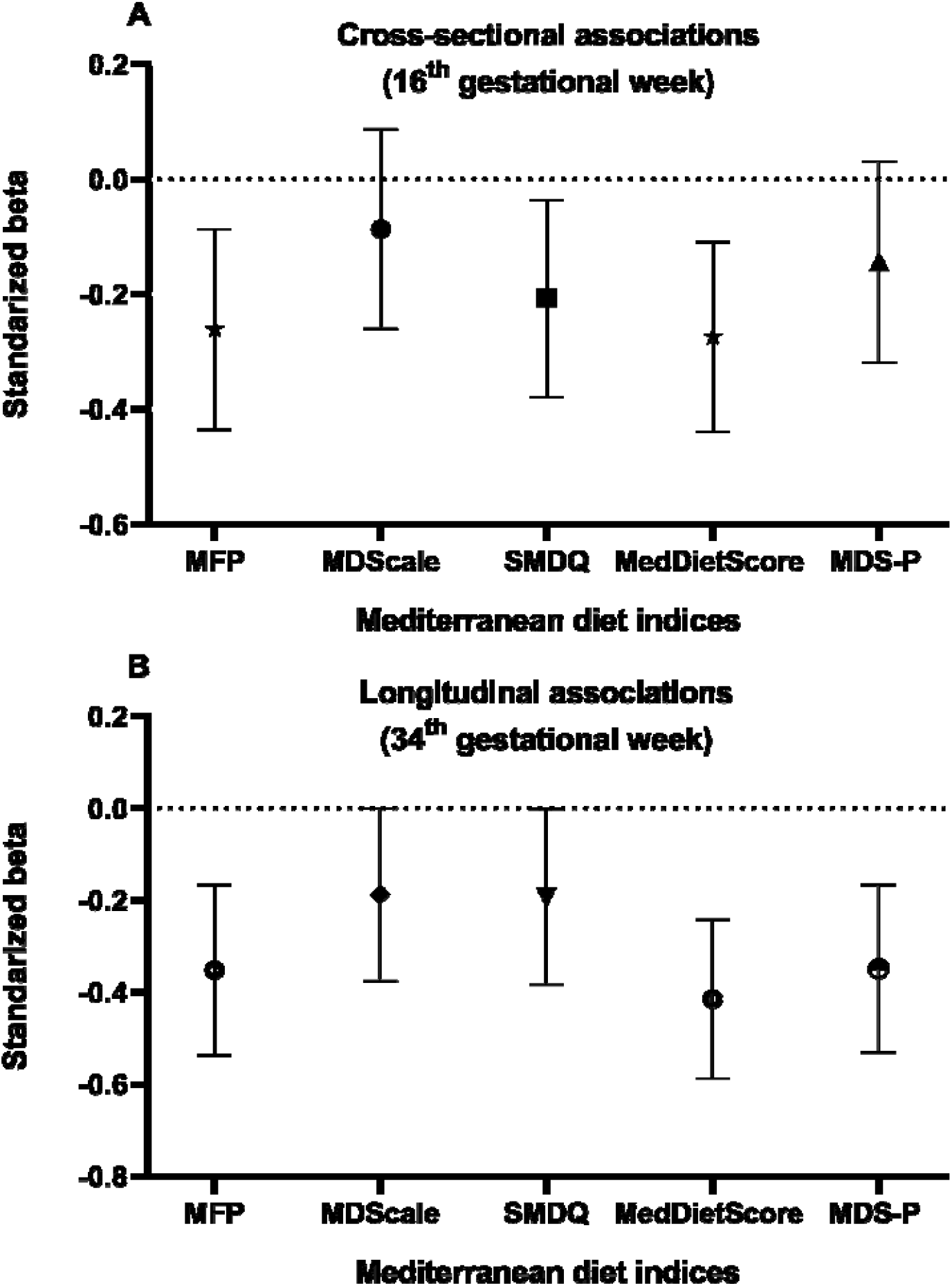
Linear regression analysis assessing the association of the Mediterranean Diet indices and the clustered cardiometabolic risk at the 16^th^ and 34^th^ gestational weeks. Dots represent β values and bars represent 95% confidence interval. (A) Cross-sectional associations of the Mediterranean Diet indices and the clustered cardiometabolic risk at the 16^th^ gestational weeks. Model adjusted for age, smoking habit and number of children. (B) Longitudinal associations of the Mediterranean Diet indices and the clustered cardiometabolic risk at the 34^th^ gestational weeks. Model adjusted for age, smoking habit, number of children and exercise intervention. MDScale, Mediterranean Diet Scale; MedDietScore, Mediterranean Diet Score; MDS-P, Mediterranean diet scale for pregnant women; MFP, Mediterranean Food Pattern; SMDQ, Short Mediterranean Diet questionnaire. Dietary indices with the same symbol did not differ significantly when compared with the J-test (*p*>0.257).

The cross-sectional and longitudinal associations between MD indices with the individual cardiometabolic markers (at the 16^th^ and 34^th^ g.w.) are shown in **Supplementary Table S3**. Higher MFP was associated with lower pre-pregnancy BMI (*p*=0.039). Greater MFP, SMDQ, MedDietScore, and MDS-P were associated with lower SBP at the 16^th^ and 34^th^ g.w. (all, *p*<0.05). Higher MedDietScore and MDS-P were associated with lower DBP at the 34^th^ g.w. (both, *p*<0.05). Greater MFP and MDS-P were associated with lower glucose at the 34^th^ g.w. (both, *p*<0.05). In addition, MFP, MDScale, and MedDietScore were associated with higher HDL-C (all, *p*<0.05).

**Table 3** and **Figure 2** show the ROC-derived thresholds together with their sensitivity, specificity, and AUC values. The MFP showed an AUC of 0.69 (95% CI: 0.59–0.79) at the 16^th^ g.w., and of 0.67 (95% CI: 0.56–0.77) at the 34^th^ g.w. The MedDietScore showed an AUC of 0.70 (95% CI: 0.60–0.81) at the 16^th^ g.w., and of 0.71 (95% CI: 0.61–0.81) at the 34^th^ g.w. The MDS-P showed an AUC of 0.62 (95% CI: 0.51–0.72) at the 16^th^ g.w., and of 0.70 (95% CI: 0.60–0.80) at the 34^th^ g.w. The cut-off points that maximized the sensitivity and specificity in each index were 21 for the MFP, 30 for the MedDietScore, and 6 for the MDS-P at both 16^th^ and 34^th^ g.w.

**Figure 3** shows the percentage of pregnant women who adhered to the MD according to the thresholds derived for the different indices recommended. Small differences (<3%) were found in the percentage of women adhering to MD across indices. The percentage of agreement and the relationship between the recommended MD indices are shown in **Supplementary Figure S2**. The lowest agreement was found between MDS-P and MedDietScore (% agreement= 67.8; kappa coefficient= 0.319), the highest was found between MFP and MedDietScore (% agreement= 76.3; kappa coefficient= 0.505).

**Figure 2.**
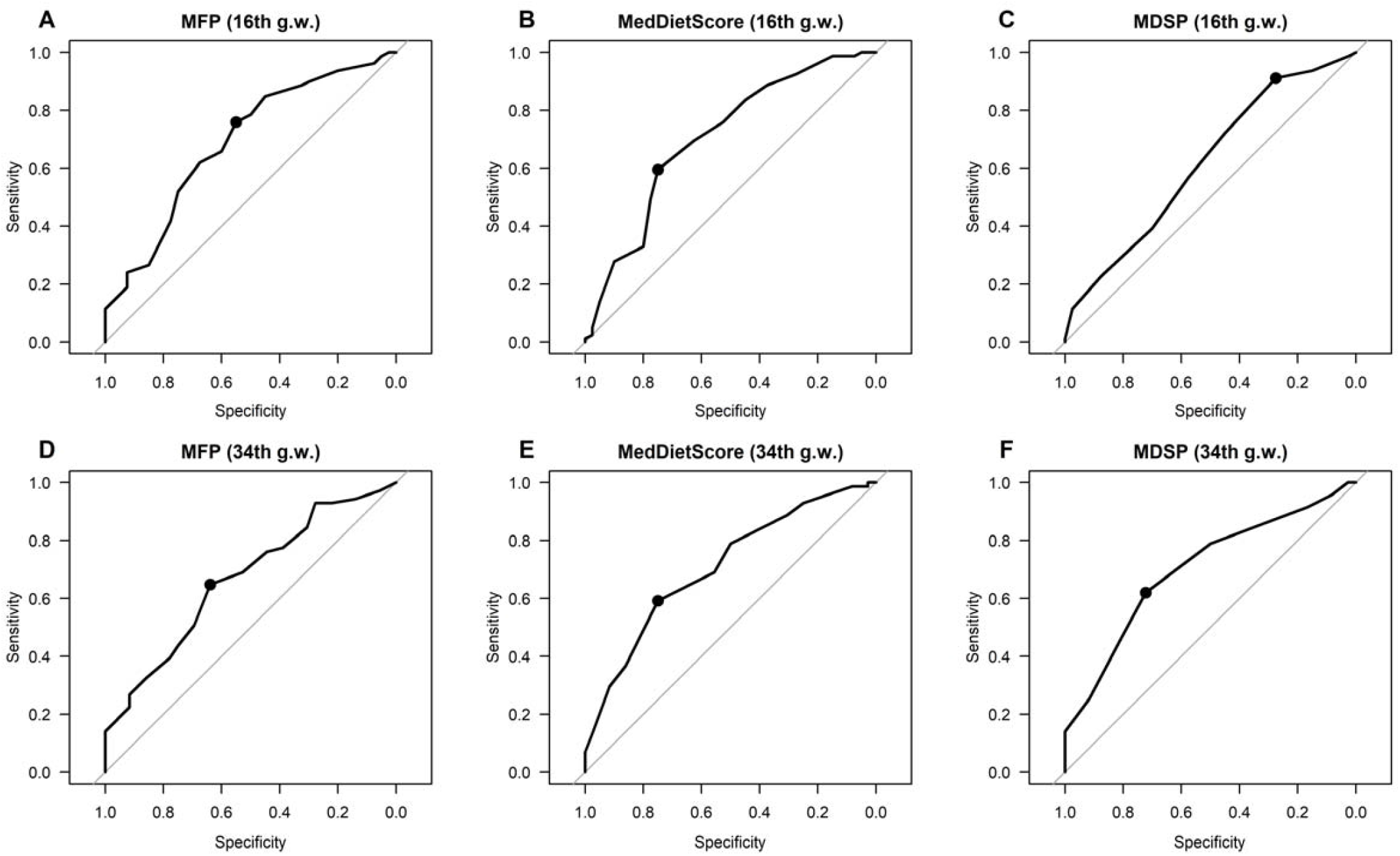
Discriminative power of the Mediterranean dietary indices recommended (ROC curves) to detect high cardiometabolic risk at the 16^th^ and 34^th^ gestational week. ROC, receiver operating characteristic.

**Figure 3.**
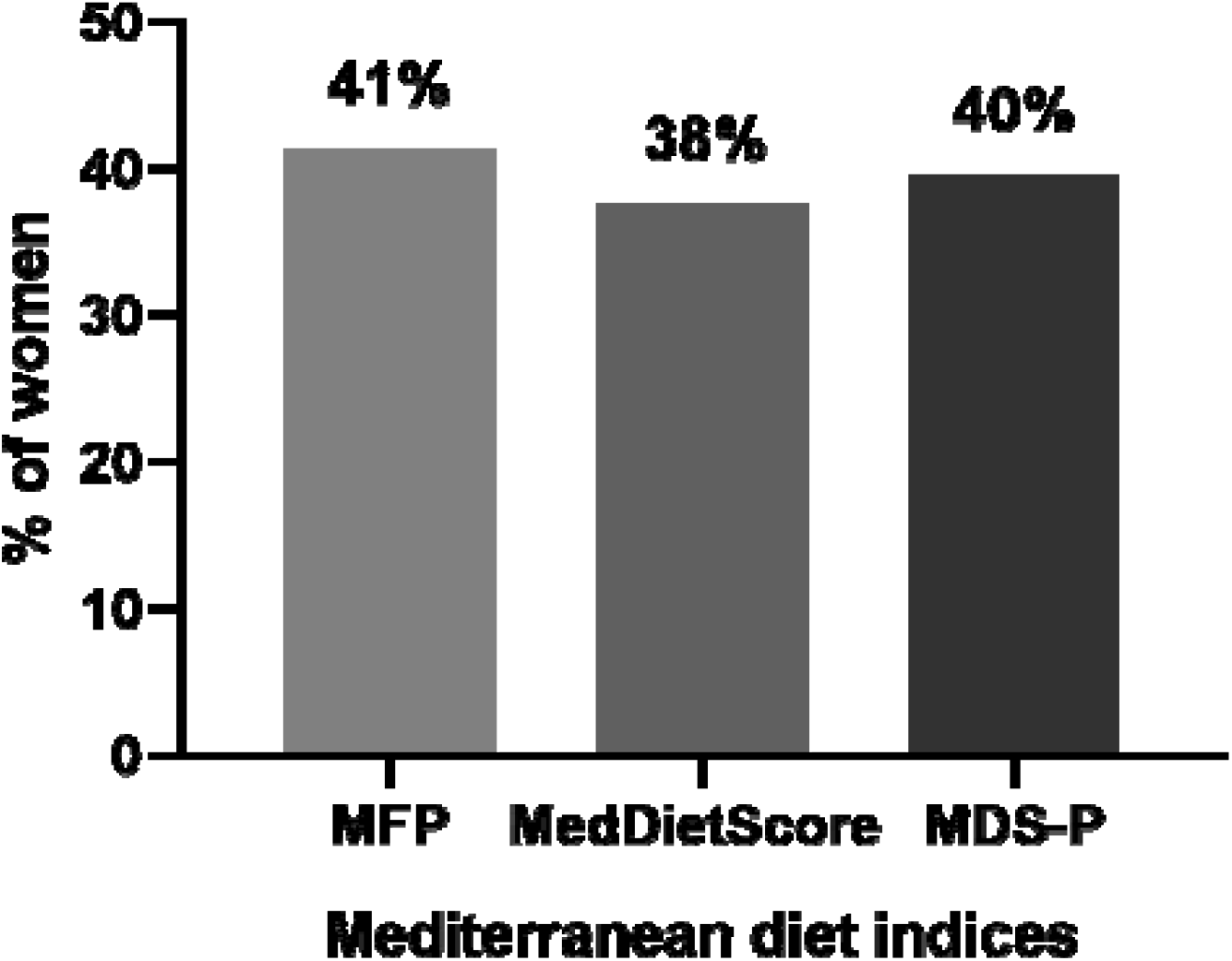
Percentage of participants who adhere to the Mediterranean diet according to the thresholds derived for the different indices recommended (n=152).

**Table 3.** Derived thresholds to identify pregnant women who adhere to the Mediterranean diet at the 16^th^ and 34^th^ gestational weeks.

| **MD indices** | **Threshold** | **Sensitivity** | **Specificity** | **AUC (95% CI)** |
| --- | --- | --- | --- | --- |
| **Clustered cardiometabolic risk at the 16<sup>th</sup> g.w.** |  |  |  |  |
| *MFP (4-35)* | 21 | 0.62 | 0.68 | 0.69 (0.59-0.79) |
| *MedDietScore (0-50)* | 30 | 0.59 | 0.75 | 0.70 (0.60-0.81) |
| *MDS-P (0-11)* | 6 | 0.57 | 0.58 | 0.62 (0.51-0.72) |
| **Clustered cardiometabolic risk at the 34<sup>th</sup> g.w.** |  |  |  |  |
| *MFP (4-35)* | 21 | 0.65 | 0.64 | 0.67 (0.56-0.77) |
| *MedDietScore (0-50)* | 30 | 0.59 | 0.75 | 0.71 (0.61-0.81) |
| *MDS-P (0-11)* | 6 | 0.62 | 0.72 | 0.70 (0.60-0.80) |

AUC: area under the curve; CI, confidence interval; G.W.: gestational week; MD: Mediterranean diet.

## DISCUSSION

Our results suggest that the MedDietScore, the MFP, and the MDS-P could be recommended to assess MD adherence during pregnancy. The MedDietScore outperformed the other indices in the association with cardiometabolic risk at the 16^th^ g.w. and 34^th^ g.w. Furthermore, the MFP was associated with cardiometabolic risk at the 16^th^ and 34^th^ g.w., and the MDS-P at the 34^th^ g.w. Among the recommended indices, the cut-off points proposed to detect high cardiometabolic risk were 21 for the MFP, 30 for the MedDietScore, and 6 for the MDS-P at both 16^th^ and 34^th^ g.w.

There is a lack of uniformity between indices to assess MD adherence during pregnancy[3,9,24], specifically in the number of components included, the classification categories, measurement scales, statistical parameters (mean, median, or quintiles of daily intake), and the positive/negative contribution of each component to the total score[39,40]. This complicates the comparability and reproducibility of the MD adherence measurement during pregnancy[3]. Previous studies have shown that maternal dietary patterns (including the MD) are associated with better cardiometabolic markers[3,41–44]. We confirmed this finding with 4 out of the 5 MD indices tested with differing association sizes and prediction capacity, suggesting that using different indices could lead to inconsistent results on the association of MD with cardiometabolic health. The MedDietScore, the MFP, and the SMDQ indices were associated with lower cardiometabolic risk at the 16^th^ g.w., and we observed that the MedDietScore and the MFP indices were superior to the SMDQ. This suggests that these two indices (i.e., MFP and MedDietScore) could be indistinctly recommended to understand the relationship between MD and the cardiometabolic risk at the 16^th^ g.w. At the 34^th^ g.w., the MDS-P was also associated with lower cardiometabolic risk. In late pregnancy (i.e., 34^th^ g.w.), the MedDietScore was superior to the MDS-P, MFP, and SMDQ indices in the association with cardiometabolic risk. Therefore, the MFP and the MedDietScore could be appropriate to assess the association between MD adherence and cardiometabolic risk at the 16^th^ and 34^th^ g.w., whereas the MDS-P could also be appropriate at the 34^th^ g.w. This recommendation is considered to be feasible since all indices could be calculated with the data obtained from the same FFQ. Discrepancies between SMDQ, MDScale and MDS-P indices regarding their association with cardiometabolic risk might be partially attributable to the components and categorization of the food components included in each index. The SMDQ index in the present study is comprised of eight elements (in servings/day or servings/week) while the MDScale includes 8 elements (in g/day) and the MDS-P additionally includes 3 elements (in mg/day or μg/day). Moreover, the MDScale relies on the median-split of the specified items and the establishment of a cut-off, in each food group, and then to the attribution of scores of 1 or 0. Otherwise, the SMDQ uses predefined cut-off portions. Regarding cereals, the MDScale includes only one cereal component (i.e., cereals) while the SMDQ index distinguishes between 3 groups of cereals such as white bread, rice), and whole grains. These components might partially explain differences between SMDQ, MDScale, and MDS-P indices and their relationship with cardiometabolic risk.

The MFP and the MedDietScore attribute a positive value when the consumption of ethanol met the established criteria. However, alcohol consumption was not considered in the scores because women must not drink alcohol during pregnancy, and data of our participants showed no consumption at all. It is worth mentioning that previous publications did not adapt the general population cut-offs points to pregnant women, which is needed given that (for example) pregnant women are not expected to drink alcohol[3,9,11]. As a result, there are no adapted cut-off points for MD adherence in pregnant women. In this study, we used ROC curves to establish such cut-off points, based on the detection of high cardiometabolic risk during pregnancy. The cut-off points that maximized the sensitivity and specificity were 21 for the MFP, 30 for the MedDietScore, and 6 for the MDS-P at both the 16^th^ and 34^th^ g.w. These cut-off points are slightly different from the previously-advised for the general population. In the original validation of the MFP index[20], it was found a negative association between the MFP score and the risk for myocardial infarction. The authors observed that the risk for myocardial infarction ceased decreasing for scores ≥20. In agreement, we found that a cut-off point of 21 in the MFP maximized both the sensitivity and specificity to detect high cardiometabolic risk. Women with more than 30 points on the MedDietScore were considered adherent to the MD. In the original questionnaire, this score corresponds to medium adherence (MedDietScore of 30-33.99) but not high adherence (MedDietScore≥34)[23,45]. Since we did not account for the alcohol intake, the maximum score considered for these analyses in the MedDietScore was 50 points instead of 55. Therefore, it seems plausible that women who are classified as adherent to the MD will have a lower absolute score compared to the one proposed in the original validation study (i.e., MedDietScore≥34). Participants with an MDS-P ≥6 were defined as being adherent to the MD in the present study, a cut-off that is considered adequate compliance (MDS-P of 5-8) but not high compliance (MDS-P≥9) to the MD according to the original questionnaire. The cut-off points in the original questionnaire were developed based on tertiles of the score distribution in the study participants and not based on a clinical outcome[24]. In order to increase the MD adherence to achieve scores greater than or equal to the ones proposed, it is necessary to increase those components that improve the MD adherence score (e.g., whole-grain cereals, fruits, vegetables, pulses or fish) and decrease those that reduce it (e.g., red meat or high glycemic foods). For instance, in the MedDietScore case increasing intake of fruit from 1-4 times/month to 13-18 times/month would increase the MedDietScore by 3 points. Similarly, decreasing intake of red meat and subproducts from 9-12 times/months to 1-4/month would increase the MedDietScore by another 2 points.

Another interesting finding was that only 40% of participants were adherent to the MD accordingly to the ROC-derived cut-off points. This concurs with recent evidence suggesting that pregnant women are drifting away from the MD[3]. Small differences (<3%) were found between the MFP, MedDietScore, and MDS-P in the percentage of individuals adherent to the MD, which increases the robustness of the finding. This could be explained because dietary indices employed in the present study were highly correlated to each other and we used consistent methods to classify the cardiometabolic risk and to derive the cut-off points for each index. This could be attributable to the fact that the same FFQ[18] was employed and the same trained nutritionist administered the FFQs, increasing the internal consistency. However, the classification agreement is far from perfect (68-76%). This indicates that around 1/3 of the women in this study are being differently classified by the indices (either in the favourable or unfavourable diet). This is also supported by the kappa coefficients, which showed poor to moderate agreement across indices. Furthermore, different indices seem to associate with different cardiometabolic risk factors, suggesting that each index might associate with a different risk profile, which might have important clinical implications. However, establishing these patterns of association is not within our study objectives, and the sample size and our study design do not allow reaching conclusive findings on this. Future studies might seek for association patterns between the indices recommended in this study and each one of the cardiometabolic risk factors.

A recent systematic review[46] where five MD assessment indices were compared, showed that the MedDietScore provides the best evidence of MD adherence. Similarly, in the present study, the MedDietScore was superior to the rest of the studied MD indices in the association with cardiometabolic risk along the pregnancy course. The MedDietScore is particularly useful because, for each of the assessed food groups, five classifications are possible; thus, it has been suggested to be more representative of the consumption of MD food items[39]. In the present study, the MedDietScore showed a high percentage of agreement with the MFP (74%) but a smaller-but still moderate-percentage of agreement with the MDS-P (68%). This concurs with previous studies[3,39] where the MedDietScore and the MDScale showed a similar percentage of agreement (i.e., 65%). MDS-P is based on the MDScale adding three components (i.e., intake of Fe, folic acid and Ca, since these micronutrients are required in optimal amounts during pregnancy). In the MDS-P one point is assigned if the participant consumes up to two-thirds of the recommended intake of Fe, folic acid, or calcium and/or takes supplements. Most participants (85%) were taking supplements which means that most of the pregnant women could have scored 3 points in those items. Therefore, not many differences may be expected between the MDScale concerning the MDS-P, explaining why a similar percentage of agreement was found between MedDietScore and MDS-P.

### Limitation and strengths

Firstly, results should be interpreted cautiously given the relatively small sample size for this specific purpose. Secondly, as this is a cross-sectional study, causality cannot be determined. Regarding strengths, we assessed dietary habits and cardiometabolic status by employing widely used MD indices which are often used as a reference in adult population studies to study cardiometabolic risk. Moreover, we included a wide range of cardiometabolic factors within the overall risk score created, which strengthens the usefulness of these proposed indices.

## CONCLUSION

We recommend the potential use of the MFP and the MedDietScore indices at the 16^th^ g.w. and the MedDietScore at the 34^th^ g.w. to assess the positive or negative cardiometabolic impact of the diet during pregnancy. ROC-derived threshold for the MFP and MedDietScore were 21 and 30 points, respectively. This suggests that these two indices could be recommended to understand the relationship between the MD and the cardiometabolic risk during pregnancy above the rest of the MD indices included in this study. This screening would help to identify pregnant women with a higher risk of complications since worse cardiovascular risk has been associated with adverse pregnancy outcomes[17].

## Supporting information

Suppementary Material

## Data Availability

The datasets used and/or analyzed during the current study are available from the corresponding author on reasonable request.

## Authorship

**Marta Flor-Alemany:** conceptualization, methodology, validation, investigation, data curation, writing-original draft preparation, writing-review, and editing. **Jairo H. Migueles:** formal analysis, writing-review, and editing. **Pedro Acosta**: validation, investigation, data curation, writing-review, and editing. **Nuria Marín-Jiménez:** validation, investigation, writing-review and editing. **Laura Baena-García:** validation, investigation, writing-review, and editing. **Virginia A Aparicio:** conceptualization, methodology, validation, resources, writing-original draft preparation, writing-review and editing, project administration, funding acquisition.

## Ethical Standards Disclosure

This study was conducted according to the guidelines laid down in the Declaration of Helsinki and all procedures involving research study participants were approved by the Ethics Committee on Clinical Research of Granada, Regional Government of Andalusia, Spain (code: GESFIT-0448-N-15). Written informed consent was obtained from all subjects.

