## Supplementary material for "Assessing the Mediterranean diet adherence during pregnancy: practical considerations based on the associations with cardiometabolic risk": Suppementary Material

**Table of contents**

**Supplementary Table S1.** Inclusion and exclusion criteria in the GESTAFIT project.

**Supplementary Figure S1.** Flow diagram of the study participants.

**Supplementary Table S2**. Components included in the Mediterranean diet adherence indices.

**Supplementary Table S3.** Association of Mediterranean diet indices with individual cardiometabolic markers during pregnancy (n=119).

**Supplementary Figure S2**. Percentage of agreement between the recommended Mediterranean dietary indices using the ROC-derived thresholds.

**Supplementary Table S1.** Inclusion and exclusion criteria in the GESTAFIT project.

**Inclusion criteria**

- Pregnant women aged 25-40 years old with a normal pregnancy course.

- Answering “no” to all questions on the PARmed-X for pregnancy.

- Being able to walk without assistance.

- Being able to read and write properly.

- Informed consent: Being capable and willing to provide written consent.

**Exclusion criteria**

- Having acute or terminal illness.

- Having malnutrition.

- Being unable to conduct tests for assessing physical fitness or exercise during pregnancy.

- Having pregnancy risk factors (such as hypertension, type 2 diabetes, etc.).

- Having a multiple pregnancy.

- Having chromosopathy or foetal malformations.

- Having uterine growth restriction.

- Having foetal death.

- Having upper or lower extremity fracture in the past 3 months.

- Suffering neuromuscular disease or presence of drugs affecting neuromuscular function.

- Being registered in another exercise program.

- Performing more than 300 minutes of at least moderate physical activity per week.

-Being engaged in another physical exercise program

- Being unwilling either to complete the study requirements or to be randomized into the control or intervention group.

Assessed for eligibility (n=384)

Excluded for different causes (n=162):

-Did not answer the phone (n=52)

-Declined to participate (n=110)

Called for first evaluation (n=222)

Excluded on exclusion criteria (n=5):

-Multiple pregnancy (n=1)

-Engaged moderate physical activity [>300 min per week] (n=2)

-Was over 16±2 weeks of gestations at the time of the first evaluation (n=1)

-Pregnancy risk factor [vaginal bleeding] (n=1)

Excluded for another cause (n=58):

-Declined to participate (n=58)

Analysed (n=159)

Analysed for sociodemographic data (**n=152**)

-Excluded from analysis (missing initial data and/or missing data in the food frequency questionnaire) (n=7)

Analysed for cardiometabolic risk data at the 16^th^ gestational week (**n=119**)

-Excluded from analysis (missing data in biochemical markers, blood pressure and/or body composition) (n=33)

Analysed for cardiometabolic risk data at the 34^th^ gestational week (**n=107**)

-Excluded from analysis (missing data in biochemical markers, blood pressure and/or body composition) (n=45)

**Final study sample (n=152)**

**Supplementary Figure S1.** Flow diagram of the study participants**.**

**Supplementary Table S2.** Components included in the Mediterranean diet adherence indices.

|  | **Food groups included in the Mediterranean diet adherence indices** | | | | |
| --- | --- | --- | --- | --- | --- |
| **Category** | **MFP** | **MDScale** | **SMDQ** | **MedDietScore** | **MDS-P** |
| **Cereals** | Cereals (g/day) | Cereals (g/day) | White bread (s/day) | Whole-grain cereals (s/month) | Cereals (g/day) |
|  | High glycemic foods (g/day) |  | White rice (s/week) |  |  |
|  |  |  | Whole grain-bread (s/week) |  |  |
| **Dairy products** |  | High fat dairy (g/day) |  | Whole dairy products (s/month) | High fat dairy (g/day) |
| **Fat** | Olive oil (g/day) | Monosaturated/  saturated lipid ratio | Olive oil (s/day) | Olive oil (s/week) | Monosaturated/  saturated lipid ratio |
| **Fiber** | Fiber (g/day) |  |  |  |  |
| **Fish** | Fish (g/day) | Fish (g/day) | Fish (s/week) | Fish (s/month) | Fish (g/day) |
| **Fruits, vegetables and nuts** | Fruits (g/day) | Fruits and nuts (g/day) | Fruits (s/day) | Fruits (s/month) | Fruits and nuts (g/day) |
|  | Vegetables (g/day) | Vegetables (g/day) | Fruits and vegetables (s/day) | Vegetables (s/month) | Vegetables (g/day) |
|  |  |  | Vegetables (s/day) |  |  |
| **Legumes** |  | Legumes (g/day) | Legumes (s/week) | Legumes (s/month) |  |
| **Meat** | Meat and subproducts (g/day) | Meat and  subproducts (g/day) | Meat (s/day) | Poultry (s/month) | Meat and subproducts (g/day) |
|  |  |  |  | Red meat and subproducts (s/month) |  |
| **Micronutrients** |  |  |  |  | Calcium (mg/day) |
|  |  |  |  |  | Folic acid (μg/day) |
|  |  |  |  |  | Iron (mg/day) |
| **Potatoes** |  |  |  | Potatoes (s/month) |  |

MDScale, Mediterranean Diet Scale; MedDietScore, Mediterranean Diet Score; MDS-P, Mediterranean diet scale for pregnant women; MFP, Mediterranean Food Pattern; S, servings; SMDQ, Short Mediterranean Diet questionnaire.

**Supplementary Table S3**. Association of Mediterranean diet indices with individual cardiometabolic markers during pregnancy.

| **Dietary Index** | **Pre-pregnancy BMI** | | **SBP** | | **DBP** | | **Glucose** | | **Triglycerides** | | **HDL-C** | |
| --- | --- | --- | --- | --- | --- | --- | --- | --- | --- | --- | --- | --- |
| **Cardiometabolic risk markers at the 16^th^ gestational week (n=119)^a^** | | | | | | | | | | | | |
|  | **Β** | ***p*** | **β** | ***p*** | **β** | ***p*** | **β** | ***p*** | **β** | ***p*** | **β** | ***p*** |
| MFP (4-35) | -0.194 | **0.039** | -0.213 | **0.029** | -0.109 | 0.264 | -0.079 | 0.413 | -0.102 | 0.289 | 0.188 | **0.040** |
| MDScale (0-8) | -0.020 | 0.830 | -0.142 | 0.135 | -0.013 | 0.890 | 0.001 | 0.995 | 0.022 | 0.817 | 0.160 | 0.069 |
| SMDQ (0-8) | -0.134 | 0.144 | -0.210 | **0.027** | -0.154 | 0.105 | 0.000 | 1.000 | -0.060 | 0.520 | 0.196 | **0.027** |
| MedDietScore (0-50) | -0.168 | 0.061 | -0.224 | **0.016** | -0.160 | 0.084 | -0.009 | 0.920 | -0.129 | 0.159 | 0.263 | **0.002** |
| MDS-P (0-11) | -0.124 | 0.180 | 0.208 | **0.029** | -0.094 | 0.326 | -0.005 | 0.956 | -0.011 | 0.905 | 0.100 | 0.266 |
| **Cardiometabolic risk markers at the 34^th^ gestational week (n=107)^b^** | | | | | | | | | | | | |
|  | **β^c^** | ***p^c^*** | **β** | ***p*** | **β** | ***p*** | **β** | ***p*** | **β** | ***p*** | **β** | ***p*** |
| MFP (4-35) | - | - | -0.215 | **0.033** | -0.167 | 0.106 | -0.210 | **0.033** | -0.124 | 0.227 | 0.254 | **0.010** |
| MDScale (0-8) | - | - | -0.201 | **0.039** | -0.092 | 0.360 | -0.177 | 0.064 | -0.037 | 0.707 | 0.160 | **0.028** |
| SMDQ (0-8) | - | - | -0.132 | 0.190 | -0.137 | 0.181 | -0.001 | 0.989 | -0.101 | 0.320 | 0.158 | 0.106 |
| MedDietScore (0-50) | - | - | -0.243 | **0.012** | -0.194 | **0.050** | -0.154 | 0.105 | -0.183 | 0.062 | 0.348 | **<0.001** |
| MDS-P (0-11) | - | - | -0.309 | **0.002** | -0.230 | **0.024** | -0.254 | **0.009** | -0.136 | 0.178 | 0.146 | 0.137 |

MDScale, Mediterranean Diet Scale; MedDietScore, Mediterranean Diet Score; MDS-P, Mediterranean diet scale for pregnant women; MFP, Mediterranean Food Pattern; SMDQ, Short Mediterranean Diet questionnaire. ^a^Model adjusted for age, smoking habit and number of children. ^b^Model additionally adjusted for exercise intervention Higher clustered cardiometabolic status entails higher cardiometabolic risk. Bold values indicate a *p*<0.05. *Since pre-pregnancy BMI is not affected by the intervention the model was not adjusted.

^c^Pre-pregnancy BMI is not shown for gestational week 34^th^ since it was measured prior to pregnancy period.

% agreement

Kappa

|  | % agreement | kappa |
| --- | --- | --- |
| MFP vs MedDietScore | 76.3158 | 0.505 |
| MFP vs MDS-P | 73.0263 | 0.44 |
| MedDietScore vs MDS-P | 67.7632 | 0.319 |

**Supplementary Figure S2.** Percentage of agreement between the recommended Mediterranean diet indices using the ROC-derived thresholds (n=152). Kappa coefficients are classified as < 0, no agreement; 0-0.19, poor agreement; 0.20-0.39, fair agreement; 0.40-0.59, moderate agreement; 0.60-0.79, substantial agreement; 0.80-0.99, almost perfect agreement; 1, perfect agreement. MDScale, Mediterranean Diet Scale; MedDietScore, Mediterranean Diet Score; MDS-P, Mediterranean diet scale for pregnant women; MFP, Mediterranean Food Pattern; SMDQ, Short Mediterranean Diet questionnaire.
